# Nanopore adaptive sampling for bacterial identification from periprosthetic joint replacement tissue

**DOI:** 10.1101/2025.03.14.25323961

**Authors:** Teresa L Street, Philip Bejon, Laura Leach, Sarah Oakley, Bernadette C Young, Nicholas D Sanderson

**Author notes:** **Corresponding author and email address Teresa Street:**. **Repositories:** Nanopore fastq data are available in the ENA under project accession: PRJEB78709. Code used for analysis is available on gitlab here: https://gitlab.com/ModernisingMedicalMicrobiology/adaptive_sampling_scripts.

## Abstract

2.

Metagenomic approaches to diagnosis of prosthetic joint infections promise more accurate and more rapid diagnosis. However, the high host DNA to bacterial DNA ratio is a challenge. Nanopore adaptive sampling (AS) can be used to preferentially sequence more of the infecting organism. Here, we evaluate AS using clinical samples from infected prosthetic joints to determine the absolute fold enrichment achieved. We found that AS achieved a range of 1.61 to 1.96-fold higher absolute fold enrichment for bacterial sequenced bases using AS over control pores. In this limited sample set, AS did not impact bacterial diagnosis overall but led to a modest increase in the bacterial sequence available without any obvious cost.

**Impact statement:** Metagenomic approaches offer the possibility to rapidly detect the cause of an infection and to provide information on drug susceptibility. Implementing this technique is challenging because samples collected from patients contain high levels of human DNA which can obscure detection of bacterial DNA. Reducing the amount of human DNA sequenced would allow easier detection of bacteria. This study assessed a sequencing protocol that rejects human DNA during the sequencing process known as adaptive sampling (AS), specifically as concerns samples from patients with joint infections. Our findings demonstrate that AS can increase bacterial sequencing efficiency. However, these modest improvements did not significantly enhance bacterial identification in our small sample set, although we did not detect additional costs associated with using AS.

The study confirms modest utility of AS in real-world clinical samples and extends current literature by applying AS to joint infection. The implications of this method extend to clinical microbiology, where rapid and accurate pathogen detection can significantly impact patient outcomes.

**Data summary:** The authors confirm all supporting data, code and protocols have been provided within the article, through supplementary data files, or in publicly accessible repositories.

Nanopore sequencing fastq data are available in the ENA under project accession: PRJEB78709.

## 5. Introduction

Direct from clinical sample sequencing has been proposed as a method to diagnose infection more rapidly than culture-based methods, and to diagnose non-culturable bacterial infection. However, clinical samples often contain high levels of host cells relative to bacterial cells, leading to the host DNA sequence swamping bacterial sequence on analysis[1]. Laboratory methods to reduce host nucleic acid contamination include differential lysis of host cells with saponin and degradation of the released DNA with nucleases. Although this is a cheap and simple depletion method achieving success [for example, 2], some studies have found this can cause bias in the bacterial composition of the sequenced sample[3] or adversely affects some species[4, 5]. Other host depletion methods include leveraging the difference in methylation between human host and pathogen DNA[6] and osmotic shock of host cells followed by degradation of released DNA by propidium monoazide intercalation[7].

The Oxford Nanopore Technologies (ONT) sequencing platform can dynamically reject nucleic acid strands from specific pores during sequencing based on set criteria. This process, known as adaptive sampling (AS), has been used as a method to reduce the amount of host DNA sequenced in a variety of clinical samples and can improve the proportion of sequence data obtained from pathogens. For example, in respiratory samples Gan *et* al observed approximately a 3.6-fold increase in *Streptococcus pneumoniae* bases sequenced from sputum and bronchoalveolar lavage fluid (BALF) samples[8] and Lin *et al* reported a 1.27 to 2.15-fold increase of human adenovirus data yield from human nasopharyngeal swabs[9]. Other sample types include vaginal swabs, where a 1.7-fold increase in bacterial sequencing depth was observed[10] and blood, where a 3 to 5-fold improvement for *Plasmodium falciparum* sequence was reported[11]. AS has also been applied to enrich sequence data for antibiotic resistance gene (ARGs), with an 8-fold increase in sequencing depth seen in BALF samples[12] and a 2-fold improvement in ARG sequencing depth from soil samples for environmental surveillance[13]. On the other hand, others have found that AS provided little or no sequence enrichment. Lin *et al* observed no enrichment of pathogen sequence data for *Chlamydia psittaci* from a BALF sample[14] although the authors acknowledge that the single sample used was likely partially degraded due to extended storage prior to extraction.

Joint replacement is a highly cost effective procedure to treat pain and restore mobility, but is complicated by infection in 1-2% of cases[15, 16]. The infecting bacteria may be present in low numbers, may be difficult to distinguish from contaminants, and traditional microbiological methods are therefore applied to multiple tissue samples taken at the time of surgery, with extended incubation periods[17, 18]. Tissue samples by definition include a large proportion of host DNA. To our knowledge, AS has not been tested for samples originating from bone and joint infections.

Adaptive sampling can be applied using two different strategies, with enriching for a specific target (enrich) such as a known pathogen reference genome or depleting a specific target (deplete) such as the human reference genome. The enrich strategy is theoretically more efficient in rejecting all reads that do not match the target but is not applicable to a metagenomic approach on clinical samples where knowledge of the target is lacking. Here, we evaluate AS with culture-positive periprosthetic tissue samples using the depletion method for contaminating host DNA removal.

## 6. Methods

### 6.1 Sample preparation and routine microbiology

Periprosthetic tissue (PPT) samples were collected intraoperatively during revision arthroplasty surgery at the Nuffield Orthopaedic Centre of Oxford University Hospitals, UK, and obtained for this evaluation following routine diagnostic workup (NHS research ethics committee reference 17/LO/1420). Seven samples were selected at random from amongst culture positive samples identified by routine microbiological assessment of periprosthetic tissue during two separate weeks in March and July 2024. Routine processing of PPT samples by the microbiology laboratory was as follows: Bactec bottles were inoculated with 0.5 ml of an inoculum generated by vortexing each tissue sample in 3 ml of 0.9% saline with sterile Ballotini balls for 15 s. Bottles were incubated under aerobic (Plus Aerobic/F culture vials) and anaerobic (Lytic/10 Anaerobic/F culture vials) conditions in a BD Bactec FX system (BD Biosciences) for up to 10 days. Any bottles that flagged positive were subcultured onto agar plates and all cultured microorganisms were identified by matrix-assisted laser desorption ionization–time of flight (MALDI-TOF) mass spectrometry on a Microflex LT using Biotyper, version 3.1 (Bruker Daltonics).

#### 6.1.1 DNA extraction

DNA was extracted from the saline inoculum, described above after vortexing with Ballotini balls. One ml was passed through a 5 µm syringe filter to remove any tissue debris. DNA was extracted from 500 µl of the filtered sample using the UCP Pathogen Mini kit (Qiagen) as per the manufacturer’s instructions, following mechanical lysis on a FastPrep-24 (MP Biomedicals), and eluted in 50 µl. Prior to preparation for sequencing, DNA was purified using 1.8x AMPure XP beads (Beckman Coulter), eluted in 20 µl molecular biology grade water and quantified on a Qubit 4.0 fluorimeter with the Quant-iT dsDNA HS Assay kit (Life Technologies). A saline-only sample (i.e. with no periprosthetic tissue) was also processed in parallel as a negative control.

#### 6.1.2 Library preparation and sequencing

Sample 1 was prepared for sequencing using the Ligation kit (SQK-LSK110, ONT) using 1 µg DNA as per the manufacturer’s instructions. 19 fmol were loaded onto a single washed v9.4.1 flow cell (with 887 pores available at the start of sequencing) and sequenced for 72 hours on a GridION MK1 with basecalling performed after sequencing using Guppy 6.3.4 and dna_r9.4.1_450bps_hac.cfg. Adaptive sampling was performed with depletion on pore numbers 1-256 using the human reference genome

(GCF_000001405.40_GRCh38.p14_genomic), with pores 257-512 used as no-depletion controls - hereafter referred to as non-AS.

Samples 2-7 did not generate sufficient DNA for preparation by ligation sequencing (400ng requirement for native barcoding ligation), so were prepared for sequencing using the Rapid PCR Barcoding kit (SQK-RPB114.24, ONT). 5 ng DNA, or where this was not possible 3 µl total volume, was used as input per sample according to the manufacturer’s instructions and including the negative control. Post-PCR, 133 ng of each sample were pooled and cleaned, and 50 fmol of the final pooled library were loaded onto a v10.4.1 flow cell and sequenced for 72 hours on a GridION MK1 with adaptive sampling performed as described above. Basecalling for these samples was carried out in real time on the GridION with Dorado and the dna_r10.4.1_e8.2_400bps_hac@v4.3.0 model.

### 6.2 Bioinformatics analysis

Basecalled fastq files were classified with kraken2[19] (v2.1.3) using a database containing human, bacterial, archaeal, and viral genomes (k2_standard_20240112, available here https://genome-idx.s3.amazonaws.com/kraken/k2_standard_20240605.tar.gz). Each read was assigned as human, target species or other, based on the highest taxonomic classification for the read. Reads were mapped to a reference genome where there were greater than 10 reads classified to a single species. Analysis scripts are available here https://gitlab.com/ModernisingMedicalMicrobiology/adaptive_sampling_scripts. Median read lengths were calculated for each sample using a subsample of up to 5000 reads. Barcode crossover effects may arise from imperfect demultiplexing and read misclassification, leading to apparent contamination where a species in another sample in the same sequencing run yields very high coverage. This was address by applying a heuristic exclusion threshold: any species with genome coverage breadth of <5% was not considered to be a true species.

## 7. Results

### 7.1 Periprosthetic tissue samples

Seven culture-positive PPT samples from seven individual patients were included in this evaluation, with one sequenced individually on a single flow cell and six sequenced with a single negative control as a multiplex on a further flow cell. All samples were culture-positive as follows: sample 1, *Enterococcus faecalis, Escherichia coli, Staphylococcus aureus*; sample 2, *E. coli*; sample 3, *E. faecalis, Proteus mirabilis, Streptococcus constellatus*; sample 4, *Enterobacter cloacae* complex, *Acinetobacter baumannii*; sample 5, *E. coli*, coagulase-negative *Staphylococcus*; sample 6, *S. aureus*; and sample 7, *S. aureus*, coagulase-negative *Staphylococcus*.

### 7.2 Sequencing results

Three samples (sample 5, 6 and 7) generated very low numbers of bacterial reads (i.e. <600), compared with 602 bacterial reads in the negative control, hence these samples were regarded as negative for the purpose of this evaluation and were not included in further analysis of AS.

Sample 1 was sequenced on a single flow and, as expected, generated more data per sample (6,874.7 Mb) than the other samples after multiplexed sequencing (median 1,117.9 Mb, interquartile range (IQR) 940.2 - 1,414.5 Mb) (Table 1). AS caused a substantial reduction in human read lengths compared to the non-AS group (Table 2). Median lengths of range 2.7 kb to 5.1 kb in the non-AS group reduced to 548 bp to 699 bp in the majority of the AS group. The exception was sample 3, where the median read length reduced from 5 kb to only 4.2 kb. However, there were only 2,652 human reads for sample 3 in the AS group, representing 0.7% of the total reads generated with AS for this sample. Overall, a reduction in median human read length enabled more bacterial bases to be sequenced in the AS group. Comparing bacterial bases generated between AS and non-AS groups shows a positive-fold enrichment across all samples when AS is applied, with between 1.61 and 1.96-fold improvements observed (Table 1).

**Table 1.** Run metrics for PPT samples successfully sequenced. Control (non-AS) and adaptive sampling (AS) group read numbers and megabases (1 million bases) shown. Absolute enrichment is the fold increase in bases for bacteria between non-AS and AS groups.

| Sample name | Multiplexed | Reads |  |  |  |  |  | Megabases |  |  |  |  |  | Absolute enrichment |
| --- | --- | --- | --- | --- | --- | --- | --- | --- | --- | --- | --- | --- | --- | --- |
|  |  | Bacteria non-AS | Bacteria AS | Human non-AS | Human AS | Other non-AS | Other AS | Bacteria non-AS | Bacteria AS | Human non-AS | Human AS | Other non-AS | Other AS |  |
| Sample 1 | No | 19,321 | 31,649 | 1,166,387 | 1,950,380 | 277,300 | 434,284 | 79.13 | 127.64 | 4,100.59 | 1,146.81 | 803.35 | 617.14 | 1.61 |
| Sample 2 | Yes | 92,598 | 179,808 | 55,546 | 111,524 | 3,276 | 6,438 | 280.83 | 541.55 | 162.8 | 101.41 | 10.55 | 20.75 | 1.93 |
| Sample 3 | Yes | 197,896 | 389,633 | 1,542 | 2,652 | 1,096 | 2,195 | 588.35 | 1,150.80 | 13.74 | 20.84 | 2.58 | 5.14 | 1.96 |
| Sample 4 | Yes | 168,747 | 329,714 | 24,577 | 47,611 | 2,013 | 4,002 | 526.62 | 1,020.02 | 75.6 | 53.31 | 5.37 | 10.51 | 1.94 |
| Negative control | Yes | 213 | 389 | 674 | 1,415 | 270 | 532 | 1.09 | 1.83 | 3.56 | 1.77 | 0.79 | 1.52 | 1.69 |

**Table 2.**
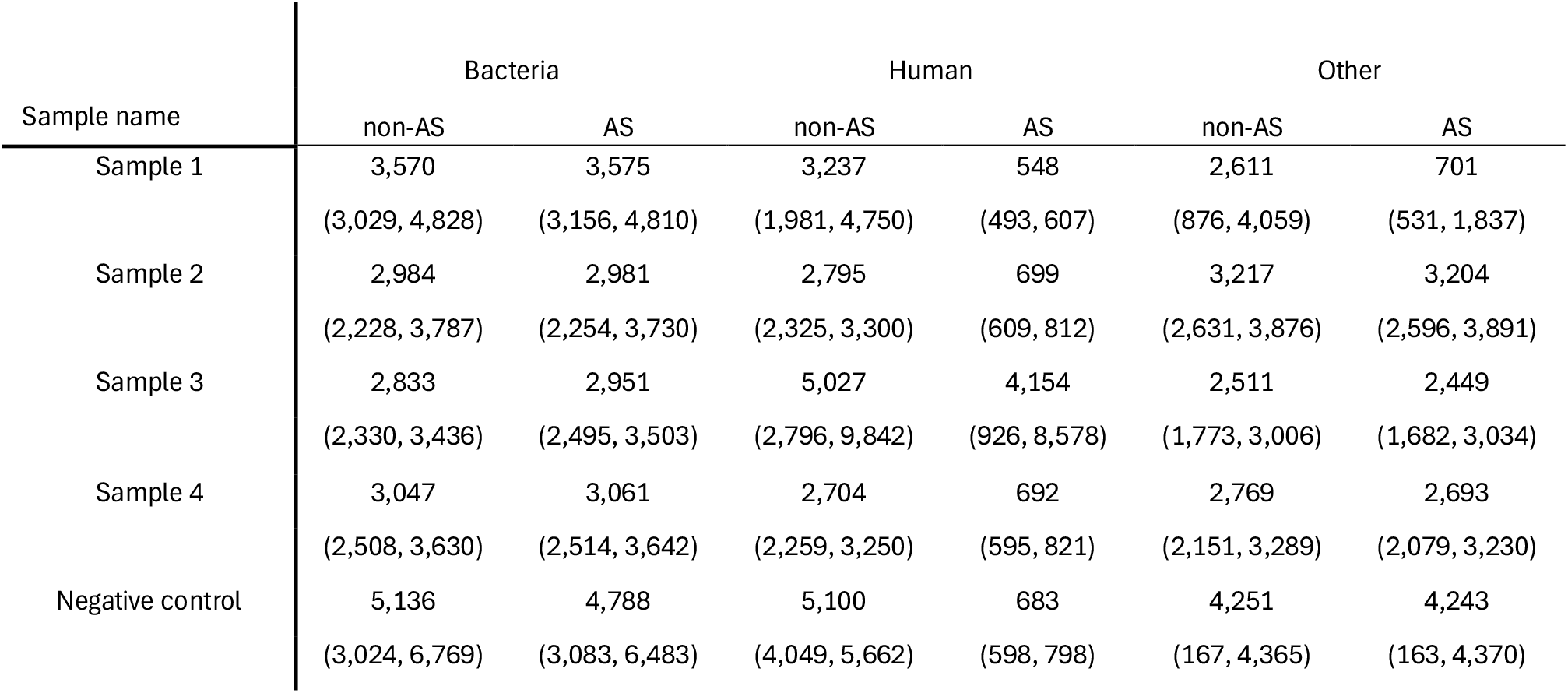
Read length median (IQR) for control (non-AS) and adaptive sampling (AS) groups, classified by kraken2 as bacteria, human or other and generated using a subsample of up to 5000 reads per taxonomic group.

| Sample name | Bacteria |  | Human |  | Other |  |
| --- | --- | --- | --- | --- | --- | --- |
|  | non-AS | AS | non-AS | AS | non-AS | AS |
| Sample 1 | 3,570 | 3,575 | 3,237 | 548 | 2,611 | 701 |
|  | (3,029, 4,828) | (3,156, 4,810) | (1,981, 4,750) | (493, 607) | (876, 4,059) | (531, 1,837) |
| Sample 2 | 2,984 | 2,981 | 2,795 | 699 | 3,217 | 3,204 |
|  | (2,228, 3,787) | (2,254, 3,730) | (2,325, 3,300) | (609, 812) | (2,631, 3,876) | (2,596, 3,891) |
| Sample 3 | 2,833 | 2,951 | 5,027 | 4,154 | 2,511 | 2,449 |
|  | (2,330, 3,436) | (2,495, 3,503) | (2,796, 9,842) | (926, 8,578) | (1,773, 3,006) | (1,682, 3,034) |
| Sample 4 | 3,047 | 3,061 | 2,704 | 692 | 2,769 | 2,693 |
|  | (2,508, 3,630) | (2,514, 3,642) | (2,259, 3,250) | (595, 821) | (2,151, 3,289) | (2,079, 3,230) |
| Negative control | 5,136 | 4,788 | 5,100 | 683 | 4,251 | 4,243 |
|  | (3,024, 6,769) | (3,083, 6,483) | (4,049, 5,662) | (598, 798) | (167, 4,365) | (163, 4,370) |

### 7.3 Bacterial species identified

The number of bacterial bases classified to the species level and aligned to reference genomes show that AS increased both the yield and genome coverage breadth for the species identified by sequencing for each sample, (Table 3, Table S1). The number of species-level bases increased from a median (IQR) of 114.4 Mb (65-298.9 Mb) in the non-AS group to 221.7 Mb (106.2-577.9 Mb) with AS.

**Table 3.** Number of mapped sequence bases and genome coverage breadth and depth split by control (non-AS) and adaptive sampling (AS) groups. Top 5 most abundant species with AS by the number of bases per sample included. Species meeting exclusion thresholds of <5% genome coverage breadth not included. Additionally, any species identified in sample culture are reported. ^a^ Percentage of the reference genomes sequenced to at least 1x depth classified by kraken2. ^b^ Non-zero average sequencing coverage depth of the sequenced reference genomes classified by kraken2, positions with no coverage are not included in the average calculation. ^c^ denotes species identified by culture for the corresponding sample. ^d^ Species detected by culture for the corresponding sample but not in the top 5 most abundant species with AS.

| Species | Mapped bases (bp) |  | Genome coverage breadth <sup>a</sup><br>(%) |  | Genome coverage depth <sup>b</sup><br>(fold) |  |
| --- | --- | --- | --- | --- | --- | --- |
|  | Non-AS | AS | non-AS | AS | non-AS | AS |
| Negative control |  |  |  |  |  |  |
| None |  |  |  |  |  |  |
| Sample 1 |  |  |  |  |  |  |
| *Enterococcus faecalis* <sup>c</sup> | 50,799,756 | 81,613,199 | 91.93 | 92.00 | 19.69 | 31.61 |
| *Escherichia coli* <sup>c</sup> | 14,245,777 | 24,448,342 | 0.14 | 0.12 | 1883.62 | 3617.69 |
| *Staphylococcus aureus* <sup>c</sup> | 39,597 | 137,715 | 1.40 | 4.64 | 1.00 | 1.05 |
| Sample 2 |  |  |  |  |  |  |
| *Escherichia coli* <sup>c</sup> | 113,588,135 | 220,371,387 | 71.34 | 71.54 | 28.96 | 56.02 |
| *Escherichia albertii* | 345,063 | 648,291 | 3.13 | 5.91 | 2.36 | 2.34 |
| *Enterobacter hormaechei* | 418,243 | 641,093 | 9.10 | 13.12 | 1.09 | 1.16 |
| *Morganella morganii* | 369,560 | 566,543 | 8.94 | 12.94 | 1.06 | 1.13 |
| Sample 3 |  |  |  |  |  |  |
| *Morganella morganii* | 450,678,041 | 881,583,379 | 93.35 | 93.35 | 124.11 | 242.79 |
| *Proteus mirabilis* <sup>c</sup> | 34,580,859 | 67,693,728 | 91.99 | 92.69 | 9.25 | 17.97 |
| *Enterobacter hormaechei* | 345,093 | 706,131 | 7.26 | 14.64 | 1.13 | 1.15 |
| *Porphyromonas asaccharolytica* | 400,823 | 656,033 | 15.88 | 23.39 | 1.15 | 1.28 |
| *Campylobacter ureolyticus* | 180,341 | 310,566 | 1.07 | 14.51 | 9.30 | 1.18 |
| *Enterococcus faecalis* <sup>d</sup> | 65,387 | 294,621 | 2.08 | 9.41 | 1.12 | 1.12 |
| Sample 4 |  |  |  |  |  |  |
| *Enterobacter hormaechei* <sup>c</sup> | 273,777,858 | 529,556,235 | 87.06 | 87.26 | 74.91 | 144.57 |
| *Enterobacter* sp. *BIDMC100* <sup>c</sup> | 11,568,088 | 22,584,779 | 20.46 | 27.42 | 12.44 | 18.12 |
| *Enterobacter cloacae* <sup>c</sup> | 7,303,047 | 13,862,857 | 43.19 | 53.52 | 3.46 | 5.30 |
| *Enterobacter cloacae* complex sp. <sup>c</sup> | 6,309,685 | 12,248,561 | 43.44 | 54.02 | 3.04 | 4.74 |
| *Enterobacter roggenkampii* <sup>c</sup> | 5,717,882 | 11,137,693 | 34.57 | 45.95 | 3.48 | 5.11 |
| *Acinetobacter baumannii* <sup>d</sup> | 472,685 | 853,225 | 10.15 | 17.91 | 1.17 | 1.20 |

The negative control sample contained reads mapping to fourteen species but these were for relatively low numbers of bases and all were removed by our exclusion thresholds (Table S1); these reads likely represent kitome or barcode crossover due to the large numbers of reads corresponding to some species seen in other samples within the multiplex, and are therefore not considered to be genuine.

Sample 1 was culture-positive for *Enterococcus faecalis, Escherichia coli* and *Staphylococcus aureus* and all three species were identified by sequencing (Table 3, Table S1). The low read numbers classified as *S. aureus* would have been interpreted as non-significant in the absence of the culture result, given the low breadth of genome covered. *E. coli* was only detected with very low breadth despite very high depth of genome coverage (Table 3); it was filtered out by our genome breadth exclusion threshold as contamination and would also have been interpreted as non-significant without culture results.

In the other polymicrobial samples, *Streptococcus constellatus* (sample 3) was detected below our mapping quality control threshold in both the AS and non-AS groups; *Streptococcus anginosus* was detected and mapped in both groups and this could represent bioinformatic misclassification due to low base numbers and similarity between species (both being members of the *Streptococcus anginous* group) (Table S1). The largest proportion of species-level reads in sample 3 were classified as *Morganella morganii* (Table 3), a species not detected by routine culture in the sample chosen for this evaluation but cultured in additional PPT samples collected concurrently from the same patient. Two additional species, *Porphyromonas asaccharolytica* and *Campylobacter ureolyticus*, were also detected in sample 3, with AS improving the genome coverage breadth of both. *Enterobacter hormaechei* detection in this sample is likely due to barcode crossover given the large number of reads classified to this species present in sample 4.

Sample 4 was dominated by *Enterobacter* reads classified as multiple separate species (Table 3), but likely representing a single species with artefactual bioinformatic misclassification. As a result of the overwhelming number of *Enterobacter* bases in both the non-AS and AS groups, *Acinetobacter baumannii*, the other species detected by culture in this sample, was detected outside the top 5 most abundant species classified. AS did, however, improve the number of bases classified and the breadth of genome coverage of this species.

In the case of a sample monomicrobial by culture, AS almost doubled the number of bases classified as *E. coli* in sample 2, which was culture positive for this species (Table 3). This improved genome coverage depth but not breadth, which was likely at the maximum coverage breadth for the automatically selected reference. In addition to *E. coli*, bases also mapped to an additional *Escherichia* species, suggestive of bioinformatic misclassification. Barcode crossover likely explains the apparent detection of *E. hormaechei* and *M. morganii*.

## 8. Discussion

Adaptive sampling, when applied to bone and joint infection, led to a modest reduction in the read lengths of human DNA without affecting bacterial DNA reads. By rejecting human reads, the aim of AS is to make sequencing pores available more frequently to sequence the organism DNA of interest. All four PPT samples with sufficient bacterial reads for analysis generated a 1.6 to 2-fold enrichment for bacterial sequence with AS compared to the non-AS control group. This is consistent with previous reports [8, 10, 12].

We generated low numbers of bacterial reads for three of seven samples. We propose this was due to a combination of factors, including the overwhelming amount of human DNA, inefficient extraction of bacterial DNA with the protocol used, and the presence of species at low bacterial loads. *Staphylococcus aureus* was only detected in sample 1 at limited genome breadth and depth and would have been interpreted as non-significant without the benefit of culture results. In addition to inefficient DNA extraction from this Gram-positive organism, there was also competition from the large number of *Enterococcus* bases sequenced in this sample. *Streptococcus constellatus* was detected at very low levels in sample 3 and this also likely represents inefficient DNA extraction given that all 5 PPT samples from this patient were culture-positive for this *Streptococcus*. Despite AS improving genome coverage breadth, *Acinetobacter baumaniii* was only detected at low levels in sample 4, which we propose was due to the very high numbers of *Enterobacter* species dominating the bacterial reads for this sample.

Sample 3 was a polymicrobial samples with 6 species cultured from the 5 PPT samples collected. The very large proportion of *M. morganii* reads detected in this sample, and the fact that two of four additional PPT samples collected from this patient cultured *M. morganii* suggests true infection, despite the sample tested here being culture-negative for this species. Additionally, the large number of *M. morganii* reads likely out competed reads for other species during sequencing, despite this sample having a relatively low fraction of human reads. In sample 4, it is likely that the mapping to multiple species reflects a single *Enterobacter* infection, with related species competing for read mapping in the workflow used.

Thresholds for species detection in metagenomic sequencing of bone and joint infection will need careful evaluation and robust thresholds will be required to assess potentially low-bacterial load samples. This will be important for addressing potential false positive and false negative results. *S. aureus* was detected just below a heuristic threshold of 5% coverage in Sample 1, and was also found in culture of that sample, as well as in one of four other PPT samples from the same patient. Previous work on the microbiology of prosthetic joint infection suggests that a single culture-positive PPT sample is poorly predictive of genuine infection, and the highest predictive value comes from finding an organism in 3 or more samples [17]. Two additional species detected in sample 3, *Porphyromonas asaccharolytica* and *Campylobacter ureolyticus*, met heuristic thresholds for this study but are unexpected species in bone and joint infection, and it is unclear if they were significant species in this context.

Sample 1 was culture-positive for *E. coli* and although this species was detected by sequencing it is more likely to represent kit contamination, or ‘kitome’[20, 21], as seen in the negative control given the very limited genome breadth observed (despite the large number of bases and depth of coverage). In this sample we see evidence that AS will amplify both true pathogens and contaminant nucleic acid.

Limitations of this evaluation include splitting the flow cell by pore number into non-AS and AS groups. A further study with greater sample numbers and controls run on separate flow cells would assess the impact of this, although variation in available pore numbers on different flow cells would also influence the interpretation of results here. Additionally, the bacterial reference genomes used were not masked for any shared sequence identity between host and target organism, and this may have reduced the specificity of the AS host read classifications resulting in erroneously unblocked reads[22]. Finally, as seen in *E. coli* culture-positive sample 2, automatic reference selection using NCBI reference genomes per species is a limitation. Highly diverse species may not achieve high genome coverage breadth due to imperfect choice of reference genome.

Adaptive sampling can increase the proportion of bacterial bases sequenced from metagenomic extractions. Whilst the fold-enrichment observed here and by others does not offer order of magnitude improvements in bacterial sequence yield, it is a simple and useful addition to a metagenomic diagnostic workflow and there does not appear to be any harm from using AS. The additional pathogen sequence data generated could theoretically contribute to the identification of low bacterial load species and enhance antimicrobial resistance determinant detection, but we were unable to demonstrate this with the limited number of samples in this study at the modest fold enrichment observed.

## Data Availability

Data are available in the ENA under project accession: PRJEB78709

## 9. Author statements

### 9.1 Author contributions

TLS, BY, PB and NDS designed the study. NDS performed the bioinformatics. TLS performed the DNA extraction and sequencing. LL and SO were responsible for sample processing and culture evaluation. TLS and NDS prepared the manuscript. All authors read and contributed to the manuscript.

### 9.4 Ethical approval

The work in this evaluation is covered under NHS research ethics committee approval (reference 17/LO/1420).

### 9.5 Consent for publication

For the purpose of Open Access, the author has applied a Creative Commons Attribution (CC BY) public copyright licence to any Author Accepted Manuscript version arising.

## 9.6 Acknowledgements

We thank the microbiology laboratory staff of the John Radcliffe Hospital, Oxford University Hospitals NHS Trust, for providing assistance with sample collection and processing.

